# Automated identification of abnormal infant movements from smart phone videos

**DOI:** 10.1101/2023.04.03.23288092

**Authors:** E. Passmore, A. L. Kwong, S. Greenstein, J. E. Olsen, A. L. Eeles, J. L. Y. Cheong, A. J. Spittle, G. Ball

## Abstract

Cerebral palsy (CP) is the most common cause of physical disability during childhood. Early diagnosis is essential to improve functional outcomes of children with CP. The General Movements Assessment (GMA) is a strong predictor of CP, but access is limited by the need for trained GMA assessors. Using 503 infant movement videos acquired at 12-18 weeks’ term-corrected age, we developed a framework to automate the GMA using smartphone videos acquired at home. We trained a deep learning model to label and track 18 key body points, implemented a custom pipeline to adjust for camera movement and infant size and trained a convolutional neural network to predict GMA. Our model achieved an area under the curve (mean ± S.D.) of 0.80 ± 0.08 in unseen test data for predicting expert GMA classification. This work highlights the potential for automated GMA screening programs for infants.

## Introduction

Cerebral palsy (CP) refers to a group of disorders that affect motor development, movement and posture and are attributed to non-progressive disturbances or injuries to the developing brain before 1 year of age^1^. Cerebral palsy is the most common cause of physical disability during childhood, occurring at a rate of 2.1 per 1000 live births worldwide^2^. While those born preterm or with low birthweight are at greater risk of having CP, almost 50% of infants with CP are born at term without overt risk factors^3,4^.

Early diagnosis is essential to improve clinical and functional outcomes of children with CP^5^. Detecting abnormal motor development within the first 6 months after birth allows targeted interventions, coincident with periods of rapid neurodevelopmental plasticity and musculoskeletal development. It has been shown that early intervention improves children’s motor and cognitive development as well parental wellbeing^5,6^. However, the average age of CP diagnosis is 19 months^3^, and only 21% of infants with CP are diagnosed before 6 months^3,7^, thus many infants miss a crucial window for early intervention.

The General Movement Assessment (GMA) can accurately predict those at highest risk of CP within the first few months after birth^8,9^. General movements are spontaneous movements involving the whole body with a changing sequence of arm, legs neck, and trunk movements^10^. Between 9 and 20 weeks of age, spontaneous movements are characterised by continuous small movements with moderate speed and variable acceleration, termed ‘fidgety’ movements^11^. These ‘fidgety movements’ are typically recognised using a trained assessor’s gestalt perception, while the infant is lying awake on their back with no direct handling or interaction^11^. This assessment is best completed from video recordings of the infant and has high predictive validity for neurodevelopmental outcomes and excellent inter-rater reliability^9,12,13^. The GMA when used during the ‘fidgety period’ has the potential to be an important screening tool in the diagnosis of CP.

The specialized training required by GMA assessors means that many primary care services and hospitals do not offer routine GMA, which limits the widespread adoption of GMA as a screening tool^16^. The ability to perform the GMA using video recordings has raised the potential to improve equitable healthcare access for those living in remote regions or in low-resource settings. Recently, the development of smartphone apps that allow the standardised recording of video by an infant’s primary carers using a hand-held device, have been shown to improve access to the GMA and allow identification of high-risk infants outside of clinical settings^12–15^. Automated scoring of the GMA from video can provide a mechanism to overcome these bottlenecks and enable high-throughput assessment for screening programs.

Recent advantages in computer vision and deep learning have led to the emergence of pose estimation techniques, a class of algorithms designed to estimate the spatial location of a set of body points from videos or pictures, and track movements with high accuracy^17–20^. Pose estimation tools do not require any specialised equipment, movement sensors or markers, and can be readily applied to track movement in new videos once trained. Several open-source pose estimation tools, pre-trained on large databases of human movement, are available for direct application to new datasets^17,21–23^. However, the standard implementation of such algorithms has been found to perform poorly in videos of infants, likely due to significant differences in body segment size and scale^24,25^. Thus, fine-tuning or re-training of pose estimation models is required to accommodate the unique characteristics of infant movement data^24^. Further, videos acquired outside of controlled, clinical or research laboratory settings may vary significantly in terms of camera angle, length, resolution, and distance to subject, requiring additional processing steps before analysis^24,26^.

Several recent studies have yielded promising results predicting motor outcomes in infants using movement tracking data from pose estimation tools^24,26–30^. Using a semi-automated approach with manual key point annotation of clinical videos, Ihlen *et al*. demonstrated computer-based movement assessments can perform comparably with observation-based GMA in predicting CP (area-under-ROC-curve, (AUC) = 0.87)^29^. Using videos acquired in a specialised laboratory setting and an infant-adapted OpenPose model, Chambers *et al*. employed a Bayesian model of joint kinematics to identify infants at high-risk for motor impairment^24^. Recent applications of deep learning models to classify movement data have also reported good performance, with one example detecting the presence or absence of fidgety movements in 5-second video clips with 88% accuracy in a laboratory setting (n=45 infants)^28^. In a large, multisite study of high-risk infants (15% with CP) Groos et al. reported a positive predictive value of 68% (negative predictive value of 95%) for later CP diagnosis using an ensemble of Graph Convolutional Networks (GCN) applied to clinical videos^27^. Similarly, Nguyen-Thai et al. applied GCNs to OpenPose tracking data to create a spatiotemporal model of infant joint movement in 235 smartphone videos, reporting an average AUC of 0.81 for the prediction of abnormal fidgety movements^26^.

Despite initial progress, significant challenges remain for the application and uptake of this technology. Many studies to-date have been limited by small sample size (typically < 100 infants) and few have been conducted outside of clinical or laboratory settings^30^. In this study, using a large cohort of infant movement videos (n=503) captured remotely via a dedicated smart phone app, we test the efficacy of automatic pose estimation and movement classification using deep learning methods to predict GMA classification. In addition, we design a custom processing pipeline to accommodate video capture from hand-held devices, identify factors that adversely affect automatic body point labelling accuracy and locate salient movement features that predict abnormal outcomes in individuals.

## Results

### Automated body point labelling with human-level accuracy

We acquired 503 3-minute videos from 341 infants at 12 to 18 weeks’ term-corrected age using Baby Moves, a dedicated smartphone app^12^. To fine-tune a pose estimation model for infant videos, we created a training dataset using a random selection of n=500 frames from 100 videos (5 frames per video, see **Methods**). We manually annotated eighteen body points (crown, eyes, chin, shoulders, elbows, wrists, hips, knees, heels and toes; **Supplemental Figure S1**) and trained a fully-customisable pose estimation model, Deep Lab Cut (DLC)^31^ to automatically detect each body point (**Figure 1** ;**Supplemental Video 1**). Body point labelling using the trained DLC model was highly accurate (**Figure 1e**) achieving an average root mean square difference (RMSD) between manual and automatic annotations of 6.78 pixels (SD: 6.60). DLC performance was comparable to inter-rater reliability (IRR) of two annotators (average ± SD RMSD 6.90 ± 7.29 pixels; **Figure 1e**).

**Figure 1:**
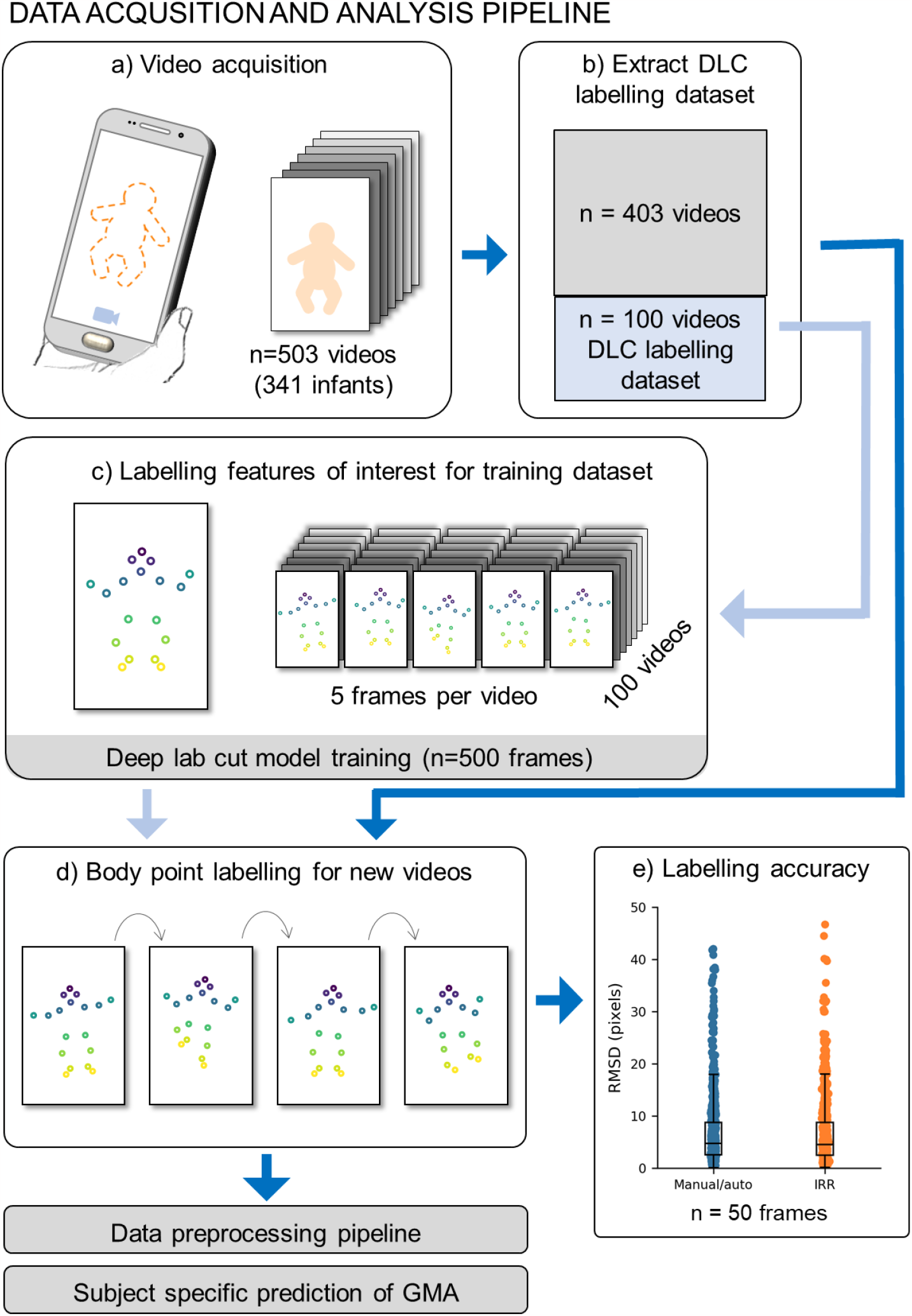
Data acquisition and analysis pipeline. **a**. Acquisition of 503 videos using the dedicated Baby Moves smartphone app^12^. **b**. 100 videos were selected for DLC training, stratified by age at video acquisition, sex and GMA classification. **c**. From each of the 100 training videos, five frames were selected for manual labelling using a k-means clustering algorithm (see **Methods**; total DLC training dataset: 500 frames). **d**. The trained DLC model was used to label all frames in all videos. This constitutes the full dataset with body point positional data used for GMA classification **e**. Labelling accuracy was evaluated in a subset of 50 frames not included in the training dataset. Root mean square difference (RMSD) in pixels was calculated between manual and automatic labelling (Manual/auto) and inter-rater reliability (IRR) of two annotators.

Labelling accuracy varied moderately across body points, with the highest accuracy for the eyes (average ± SD RMSD: manual/auto Left 3.04 ± 2.24 pixels, right 3.61 ± 2.63 pixels; IRR Left 1.80 ± 0.86 pixels, right 2.32 ± 1.22 pixels) and lowest accuracy for the hips (average ± SD RMSD: manual/auto Left 10.37 ± 6.13 pixels, right 12.02 ± 8.41 pixels; IRR Left 10.05 ± 7.20 pixels, right 14.14 ± 8.85 pixels) (**Supplemental Figure S3**). There was no significant difference in RMSD between different video resolutions (**Supplemental Table S2**).

During labelling, the DLC model assigned each point a measure of prediction confidence. After removing points labelled with low confidence (see **Methods**), we found that the percentage of frames labelled on average was 92% (SD: 16%). The percentage of frames in which each point was confidently labelled was lowest for the wrists (average ± SD Left: 81 ± 21%, Right 78 ± 24%) and heels (average ± SD: Left: 69 ± 24%, Right 77 ± 20%) (**Supplemental Figure S4**), due in part to these body points being occluded by other body parts at instances throughout the video and exhibiting a greater range of movement. We conducted a sensitivity analysis to determine potential factors that related to labelling failures (see **Methods**). We found that the amount of clothing worn by the infant moderately affected model performance with outfits that covered the hands and feet adversely affecting labelling of the extremities (F=5.180, p=0.006; **Supplementary Table S4**). As a quality control step, only videos where on average at least 70% of body points per frame were confidently labelled were included for further analysis. After quality control, our final cohort comprised n=484 videos from 327 unique infants.

### Predicting GMA from video data

As videos were not acquired in standardised clinical or experimental settings, positional data were pre-processed using a custom pipeline to account for different video acquisition parameters and potential camera movements relative to the subject, prior to classification. This consisted of outlier removal, gap filling, adjusting for camera movement, scaling to unit length based on infant size and framerate normalisation (see **Methods**). After pre-processing, each video was represented as a 46 × 4500 feature-by-frame matrix comprising standardised *x* and *y* coordinates of each body point and 2D joint angles of 10 joints in each frame.

Abnormal movements may occur at any point during the video and occur with different frequencies, therefore we aimed to identify short periods where discriminant movements were present using a sliding window approach (**Figure 2**). We trained a convolutional neural network to predict GMA based on short instances of positional data over time (**Figure 2c**), calculating video-level predictions by integrating over all clips for a given video.

**Figure 2:**
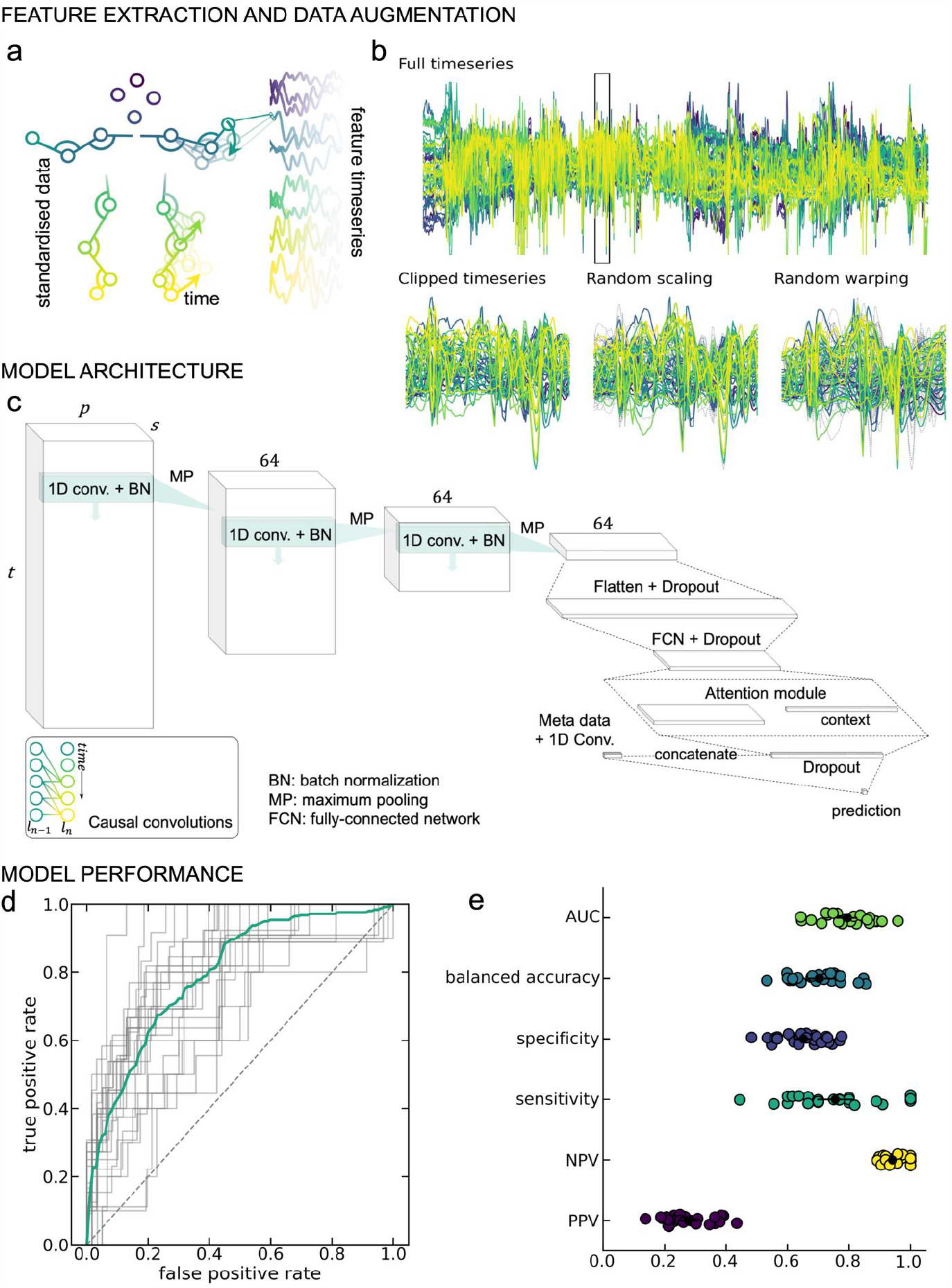
GMA prediction from movement data. **a**. Framewise positional data from DLC labelled videos were preprocessed to derive a set of feature timeseries (46 features × 4500 frames) per video. **b**. The classification model was trained on 128-frame clips for the full timeseries (top). Data augmentation steps (magnitude scaling and time warping; bottom middle and right) were applied to each clip during training. For each augmentation method, dashed grey lines indicate timeseries position prior to the augmentation step. **c**. Model architecture. 1D convolutional layers were combined with an attention module to classify normal and abnormal/absent GMA. Causal convolutions (inset) were applied to account for the temporal structure of the data. **d**. Receiver-operator curves (ROC) for each of the 25 cross-validation repeats. The mean curve is overlaid in teal. **e**. Model performance statistics for each of the cross-validation repeats. Mean and standard deviation across repeats are overlaid in black. AUC = area under the ROC; NPV = negative predictive value; PPV = positive predictive value.

Averaged over 25 cross-validation repeats (70% train/15% validation/15% test), the trained model achieved an AUC (mean ± S.D.) of 0.795 ± 0.080 in unseen test data (**Figure 2d**) and balanced accuracy of 0.703 ± 0.083 (**Figure 2e**). For abnormal/absent GMA, the positive predictive value (PPV) was 0.277 ± 0.077 and the negative predictive value (NPV) was 0.941 ± 0.035. Sensitivity and specificity were 0.755 ± 0.150 and 0.651 ± 0.078, respectively (**Figure 2e**).

Performance was consistent over a range of model parameters, including batch size, learning rate and weight regularisation (**Supplementary Figure S5**). The inclusion of video metadata, age at video acquisition and birth cohort (extremely preterm or term-born infants) improved model performance significantly (**Supplementary Figure S5**), compared with classification using video data alone (AUC = 0.749 ± 0.077). Taking advantage of multiple model instances trained across different cross-validation repeats, we found that individual predictions were generally stable when videos were included in the held-out test set for a given repeat (**Figure 2d-e, Figure 3**Error! R eference source not found.).

**Figure 3:**
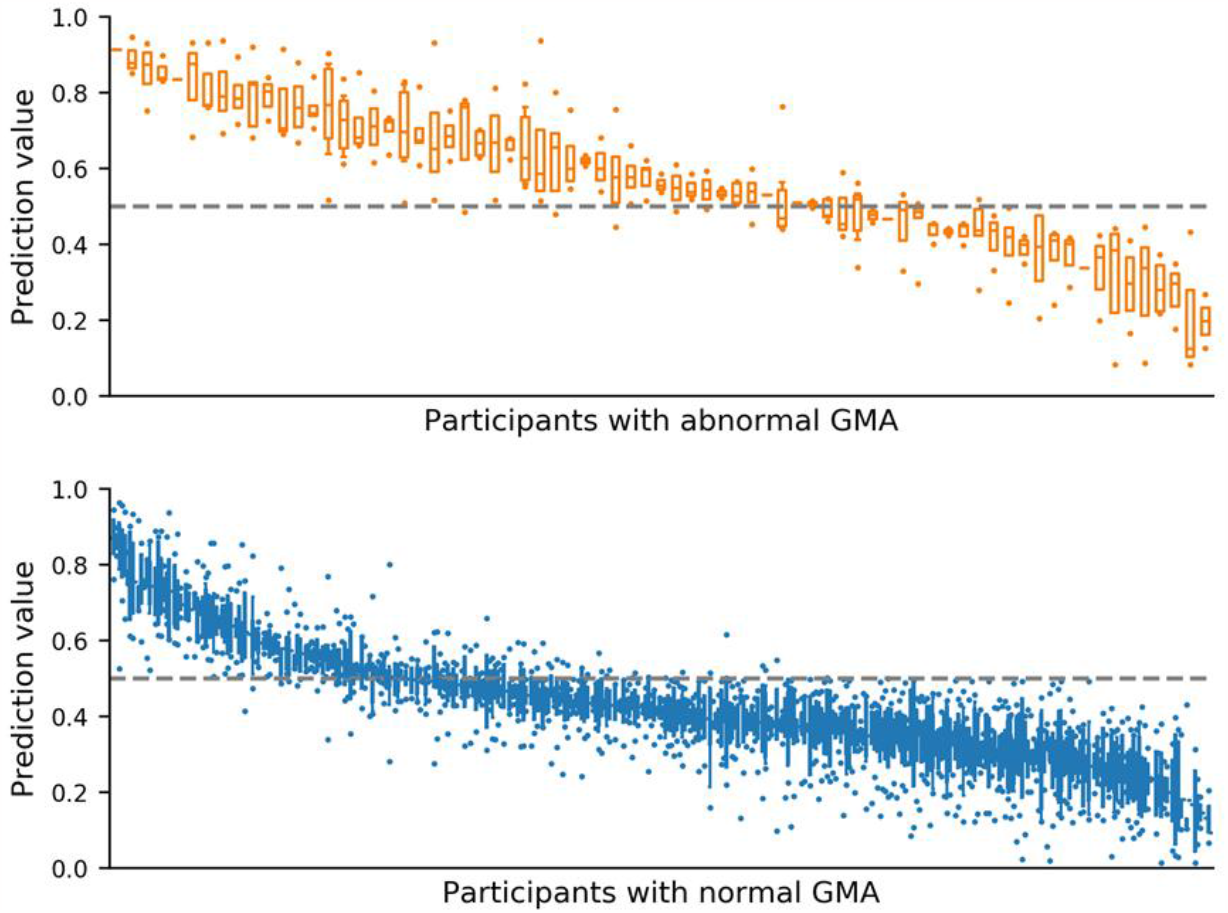
Variation in classifier prediction values. Classifier prediction values from 25-fold cross validation. Prediction values reported when infant movement included in held out test set, ordered by median prediction value. Top (orange) infant videos scored as abnormal/absent general movement assessment (GMA) by expert rater, n=73 infant videos. Bottom (blue) infant videos scored as normal GMA by expert rater, n=396 infant videos. Boxes with horizontal line represent interquartile range and median respectively, error bars represent 95% confidence interval and dots represent outliers. Dashed line at 0.5 represents cut-off value for classifier between abnormal/absent GMA prediction and normal GMA prediction.

We compared performance with an alternative baseline model: an *l*_2_-regularised logistic regression applied to a set of timeseries features extracted from each video^32^. The baseline model achieved an AUC of 0.706 ± 0.098 (0.604 ± 0.106 without video meta-data). A nonlinear, kernelized logistic regression model achieved cross-validated AUC = 0.720 ± 0.100.

### Examining spatial and temporal model attention during prediction

To identify potential features that were important to model prediction, we computed spatiotemporal saliency maps for each video^33^, (**Figure 4a**). This value highlights features (for a given body point in a single video frame) where changes in input would elicit the largest change in model prediction and can be used as a measure of model sensitivity to input data^33,34^. An example saliency map is shown for a single subject in **Figure 4a**. Saliency varied across the length of the video, corresponding with variations in model attention (white line, **Figure 4a** bottom). Clips with high saliency, relative to all subjects in the test set, are highlighted with yellow bars on the input feature timeseries, illustrating model attention to periods of different length spread throughout the video. Averaging total saliency across all clips for each body point reveals higher model sensitivity to position of the lower body points (**Figure 4a** middle), including movement of the knee and ankle joints.

**Figure 4:**
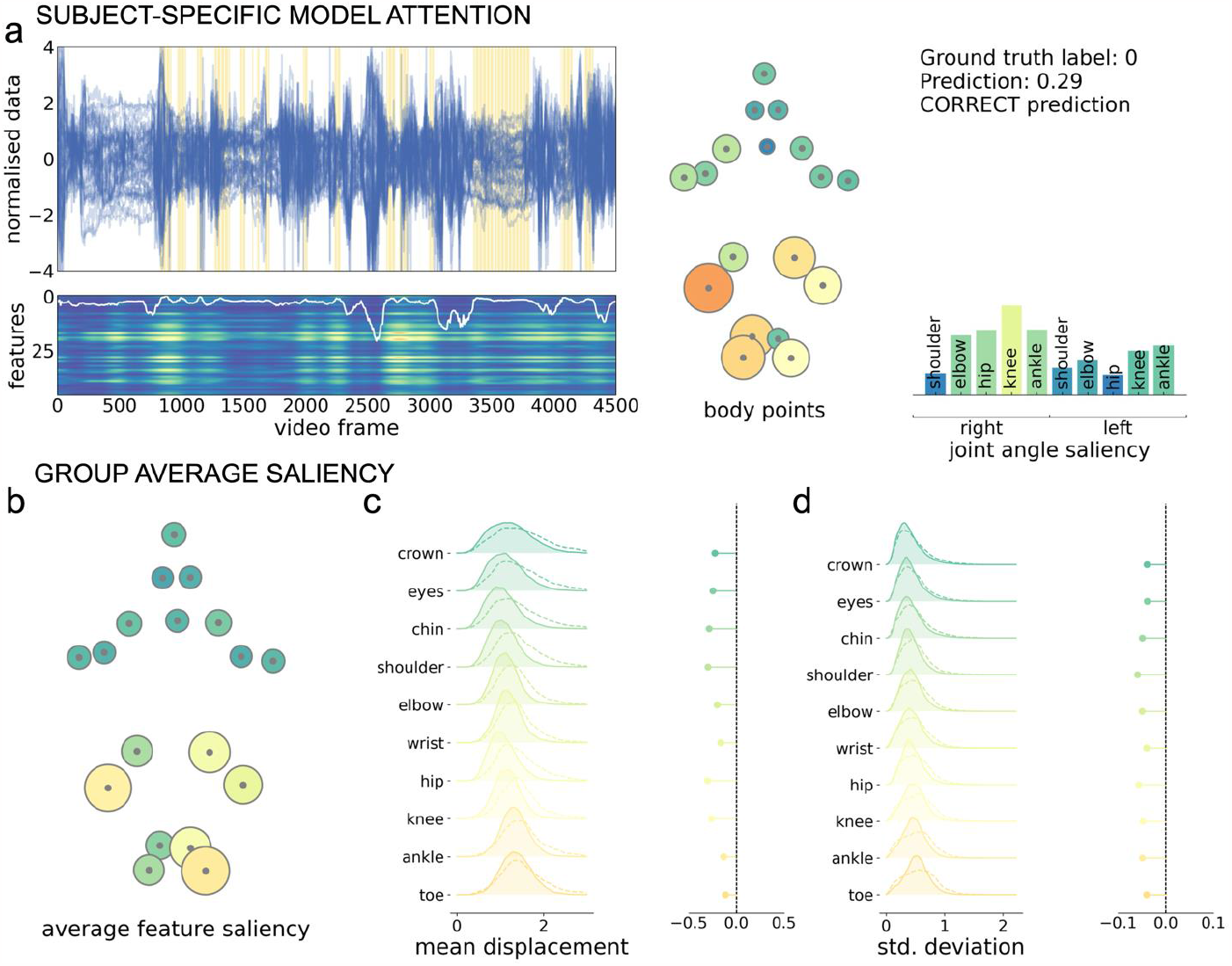
Sensitivity of model predictions to input features. **a**. Feature timeseries (top) and saliency map (bottom) for a single, correctly-classified video from an infant with normal GMA. Timeseries are shown for each feature (n=46), across the length of the video. Yellow bars indicate clips with high model attention (75^th^ percentile across all subjects). Saliency was calculated for each feature in each frame and summed over frames within each clip (n=547 clips). The map has been upsampled and smoothed to match the length of the timeseries (frames=4500). Lighter colours indicate higher saliency (arbitrary unit). Clip attention derived from the attention module (upsampled and smoothed) is overlaid in white. Average saliency across the full video is shown for each body point (middle) and joint angle (right). The model prediction is shown top right, where 0 indicated normal GMA prediction. **b**. Body point saliency averaged across all participant videos. Lighter colours and larger size reflect higher saliency. **c**. Left, mean absolute distances between each body point and their respective average position during clips of high (solid line, filled) and low (dashed line) saliency. Density plots show the distribution of displacements for high and low clips over all videos and cross-validation repeats. Right, median difference between displacement in high and low clips. **d**. Left, standard deviation (std) of displacements from the average position in high and low saliency clips, over all videos and cross-validation repeats. Right, median difference in standard deviation distributions.

Similar patterns of model saliency were observed across all participants. A map of group average feature saliency (averaged across clips, participants and cross-validation repeats) is shown in **Figure 4b**. Model saliency was highest in the lower body. This pattern was consistent across cross-validation repeats (**Supplemental Figure S*7***) and between normal and abnormal/absent GMA predictions (**Supplemental Figure S6**).

To further characterise features to which the model prediction was sensitive, we compared timeseries data in clips with high (90^th^ percentile) and low (10^th^ percentile) total saliency (**Figure 4c-d**). The number of high saliency clips did not differ between normal (mean ± S.D. = 55.21 ± 33.93) and abnormal/absent (51.74 ± 35.56) GMA videos (**Supplemental Figure S8**). For each clip, we calculated the mean (absolute) displacement of body points from the average position, as well as the standard deviation of displacements over frames. We found that, high saliency clips were characterised by body point positions closer to the average body position (**Figure 4c**) and by a lower standard deviation of joint displacements over time compared to clips with low saliency (**Figure 4d**).

### GMA prediction and development at 2 years

We compared our model predictions with participant’s motor, cognitive and language outcomes at 2-years corrected age as assessed by the Bayley Scales of Infant and Toddler Development-3^rd^ edition (Bayley-III) (**Figure 5**; **Supplemental Figure S9, Figure S10**). We found strong evidence for differences in 2-year motor composite scores between infants with different predicted GMA classifications (normal vs abnormal) when metadata (age at acquisition and birth cohort; extremely preterm or term-born infants) were included in the model, mean difference 11.35 (95%CI = [8.41, 14.30], t(439)=7.574, p<0.001). These differences were diminished when using movement data alone, mean difference 2.33 (95%CI = [-0.88, 5.54], t(439)=1.426, p=0.1555) (**Figure 5a**). There was also strong evidence for differences in 2-year cognition and language composite scores between predicted GMA classifications (**Supplemental Figure S9, Table S4**).

**Figure 5:**
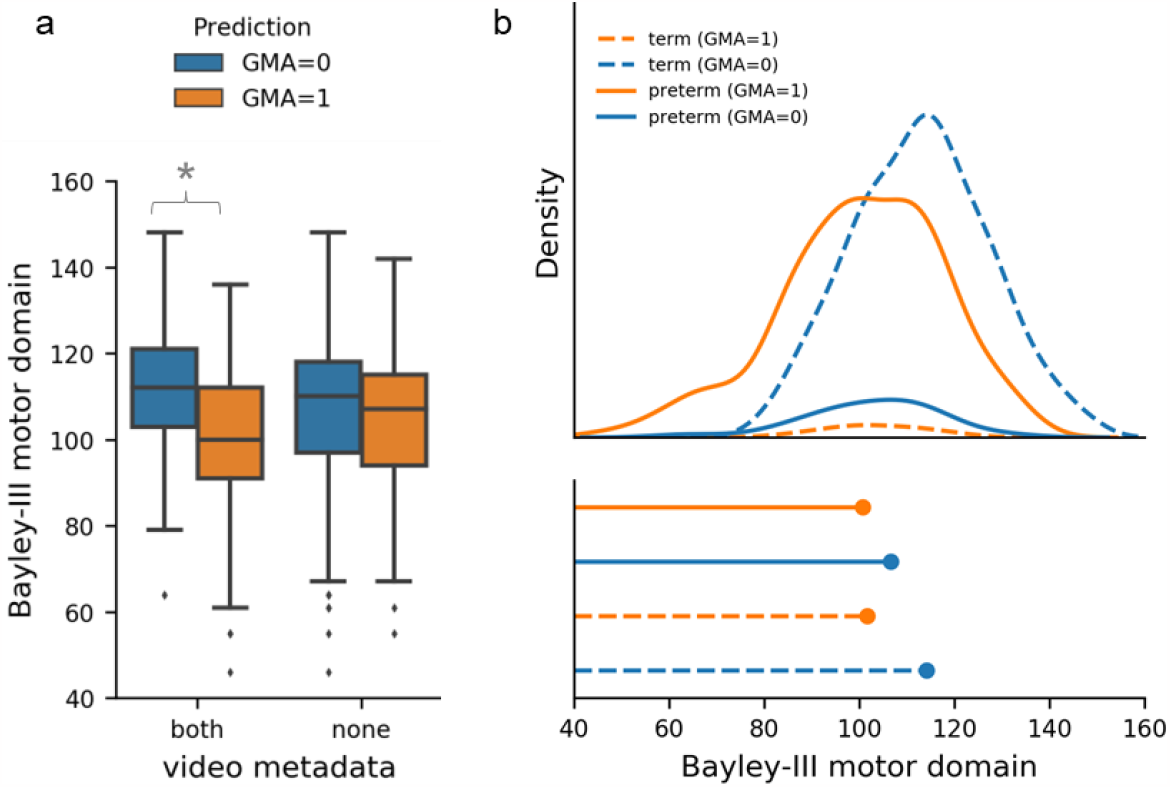
GMA prediction and motor development at 2 years: **a**. Bayley-III motor outcome stratified by GMA prediction (n=441 infant videos) and model variants trained using video movement and metadata (both = age at acquisition and birth cohort) and movement data alone (none). Blue indicates GMA prediction = 0 (normal) and orange indicates GMA prediction = 1 (abnormal/absent) for all graphs. * Indicates strong evidence for differences between GMA prediction groups from independent two-sample t-test. **b**. Top, density function of motor outcome by birth cohort and GMA prediction. Bottom, Peak of density function. Term infants are represented by dashed lines and preterm infants by solid lines.

As preterm birth is associated with both higher risk of abnormal GMA and poor neurodevelopmental outcomes, we conducted a secondary analysis to identify associations between birth cohort, GMA prediction and 2-year outcomes. We found a significant main effect of birth cohort (F=8.775, p=0.003) but not GMA prediction (F=1.762, p=0.185) on motor composite scores. The interaction between birth group and GMA prediction was not significant (F=1.181, p=0.278). Stratifying by birth cohort, there was weak evidence for differences in motor composite scores between GMA classification categories in term born infants, mean difference 9.52 (95%CI = [-1.16, 20.20], t(225)=1.756, p=0.080). In preterm infants, these differences were lower 2.09 (95%CI: = [-5.11, 9.29], t(212)=0.573, p=0.568) (Figure 5b). Similar results were observed in cognitive and language composite scores (**Supplemental Figure S10, Table S6**).

## Discussion

Using deep learning applied to smart phone videos, we tracked infant movements at 12-18 weeks of age, predicting GMA ratings outside of a controlled clinical setting. Our paper illustrates the potential for early automated detection of abnormal infant movements implemented through at-home video acquisition.

Our best performing model for predicting expert GMA ratings, was a deep learning model, consisting of 1D convolutions and an attention module. Our model achieved an AUC 0.80 (SD: 0.08), comparable to results obtained from Ihlen et al. and Groos et al. in cohorts of high-risk infants using video recordings from stationary cameras in clinical settings^27,29^. Our model outperformed alternative baseline models and was robust over various hyperparameter settings. We demonstrated that including participant metadata is crucial to improving model predictions, highlighting the increased risk for abnormal movements in preterm born individuals^8^. Of the 41 infants with abnormal GMA as scored by trained GMA assessors 35 (85%) were from preterm infants and 6 (15%) from term-born infants. We found that including metadata (birth cohort and age) improved model performance from an AUC=0.70 based on movement data alone to AUC=0.80. Notably, classifying on birth cohort alone would result in an AUC of 0.69, quantifying the added value of the movement data.

GMA model predictions were associated with poorer neurodevelopmental outcome at 2 years of age. We found that this effect was largely dependent on preterm birth, although infants with abnormal GMA predictions scored lower on average regardless of birth cohort. The association between preterm birth and poor neurodevelopmental outcomes is well established^8,13^ and this finding reflects the relatively lower predictive validity of GMA ratings, and therefore model predictions, for motor and cognitive outcomes at 2 years.

GMA is a strong predictor of CP^8,9^. We used abnormal or absent GMA ratings as a surrogate measure for CP risk. Fidgety movements can be classified as abnormal or absent during this developmental window, both of which are associated with neurodevelopmental impairment^8,13^. Combining abnormal and absent groups, who may have different movement signatures, into a single cohort may have affected model performance but numbers were too small to further split the groups (n=40 and 36 respectively). Similarly, only 6 infants in the current cohort were diagnosed with CP by 2-year follow-up, precluding the use of our model framework to predict CP diagnosis in this group.

To track infant movement, we used a pose-estimation algorithm, Deep Lab Cut^31,35^, which has the advantage of being customisable across species, age and features of interest using a minimal training dataset^35^. Our model achieved human-level labelling accuracy of body parts with a RMSD of 6.78 pixels (SD: 6.60). Due to the non-controlled settings in which videos were acquired, we performed a sensitivity analysis, identifying video features that could affect labelling accuracy in at-home video recordings. Body point labelling was robust to background and video lighting but moderately affected by clothing worn by the infant, specifically clothing covering hands or feet. Use of at home video recordings introduced additional data processing challenges, including camera movement relative to the infant and different video formats (frame rate, resolution, distance from infant). To accommodate this, we developed a novel framework, that is versatile and supports videos taken outside a standardised clinical setting. Future work could focus on more detailed anatomical annotations for labelling, particularly relating to the hands and feet in recognition of the role those body parts play in identification of fidgety movement during the GMA^11^.

We used 5 second clips to train our model, this allowed us to identify periods of movement that were informative to the model prediction. By analysing model saliency, we were able to extract information about which movement features were attended to by the model, discriminating between abnormal and normal movements. We identified that lower limb movements contributed more to the classifier’s output, with higher saliency attributed to video clips where infant position was closer to the average position, ignoring periods where the infant has moved significantly from the supine position (i.e.: out-of-frame movement, rolling). Other studies have used high-resolution video annotations identifying periods of abnormal movement within videos^29^. While this approach is likely to improve automated movement identification it requires a significant amount manual annotation and labelling that would be difficult to achieve in larger cohorts.

Several recent studies have yielded promising results predicting motor outcomes in infants. However, to date these studies have been limited by small sample size (typically < 100 infants), only including high-risk infants and few have been conducted outside of clinical or laboratory settings^30^. A strength of our study is the inclusion of both extremely preterm and term born infants within our dataset. Our study offers an automated approach to perform GMA ratings, that is capable of accommodating videos recording outside the clinical setting. Our work highlights the potential for automated approaches to screen for CP at a population level, which would enable increased access to early interventions for these children.

## Methods

### Participant data

Videos were recorded using the Baby Moves smart phone app by the parent/caregiver on their personal device between April 2016 and May 2017^12^. Videos were acquired from n=155 (77 female [50%]) extremely preterm infants (<28 weeks’ gestation) and 186 (91 female [49%]) term-born control infants, **Supplementary Table S1**. In total, 503 videos from n=341 infants aged between 12- and 18-weeks term corrected age were available. For a subset of n=160 (75 preterm, 85 term), two videos were collected per infant during this period. Full details of the study protocol can be found in Spittle et al., (2016)^12^. The study was approved by the Royal Children’s Hospital Ethics Committee (HREC35237).

### Video capture

To facilitate video recording outside of clinical or laboratory settings, the Baby Moves app provides detailed instructions and a dotted outline overlay to improve positioning of the infant in the video frame^12^. Guidance was given to parents/caregivers to perform the video while the infant was lying quietly and not fussing with minimal clothing, consisting of singlet and nappy only. Subsequently, videos were securely uploaded to a REDCap database^36,37^ at the Murdoch Children’s Research Institute for remote review. The GMA was scored according to Prechtl’s GMA^11^ by two independent assessors that were unaware of participants’ neonatal history. General movements were classified as normal if fidgety GMs were intermittently or continuously present, absent if fidgety GMs were not observed or were sporadically present, or abnormal if fidgety GMs were exaggerated in speed and amplitude. If there was disagreement between the two assessors, then a third experienced GMA trainer and assessor made the final decision. Any videos rated as unscorable were not evaluated in this study.

All videos were submitted in MP4 format. Due to differences in device model and settings, three video resolutions were present in the dataset: 480 × 360 (n=366 videos), 640 × 480 (13 videos) and 720 × 480 (126 videos) with a median frame rate of 30 frames per second (range: 15 to 31). Each video was 3 minutes in length resulting in mean (SD) 5100 (497) frames per video.

### Automated body point labelling

We trained a deep learning model using Deep Lab Cut, version 2.1 (DLC)^31^ to label and track key body points. To train the DLC model, we formed a training dataset consisting of a subset of 100 videos from our dataset stratified for age, sex, birth cohort (preterm or term) and video resolution. Only one video per infant was allowed in the training set. For the training dataset, five frames from each video were manually labelled with 18 key body points: crown, chin, eyes, shoulders, elbows, wrists, hips, knees, heels and big toes (**Figure 1**; **Supplemental Figure S1**). Manual labelling was performed via the DLC graphical user interface. To ensure diversity of movements in the frames selected for labelling, we used a k-means clustering algorithm implemented in DLC to select five frames from different clusters within each video for labelling. We implemented a DLC model with a pre-trained ResNet-50 backbone and trained for 1 million iterations on a NVIDIA TITAN Xp using a training/validation fraction of 0.95/0.05 (see **Supplemental Figure S2** for training performance).

Once trained, the DLC model was used to automatically label the 18 body points for all videos in the dataset. For each frame, the DLC model returned the x- and y-coordinates in pixels of the body points relative to the corner of the video image and its prediction confidence. Body points with a prediction confidence below 0.2 were removed. Labelling accuracy of the DLC model was evaluated on an independent sample of 50 random frames not included in the training dataset using the root mean square difference (RMSD) between predicted labels and manual labels. To evaluate inter-rater reliability (IRR) for body point labelling a second human annotator repeated labelling on the same 50 frames. The RMSD between labels for the two annotators were calculated. Additional metrics of DLC model performance included the number of unlabelled body points per video. As videos were collected outside of a controlled clinical setting, we conducted a sensitivity analysis to determine whether variability in certain factors across the individual videos may influence model performance. Each video not included in the training dataset (n=403) was categorised by the following factors: Lighting (Dark/Okay/Bright), clothing (Bodysuit/Nappy & singlet/Nappy only), skin tone (Light/Fair/Medium/Dark), infant in frame entire video (Yes/No), background (Pattern/Solid Colour – Dark/Solid Colour – Light), extra items in view (No/Another Child/Recorder’s feet/Toys/Other). We tested if model performance was affected by the listed factors using a mixed model Analysis of Variance (ANOVA).

### Pre-processing pipeline

Data pre-processing consisted of quality control, outlier removal, gap filling, adjustment for camera movement, scaling and feature extraction (**Figure 6**).

**Figure 6:**
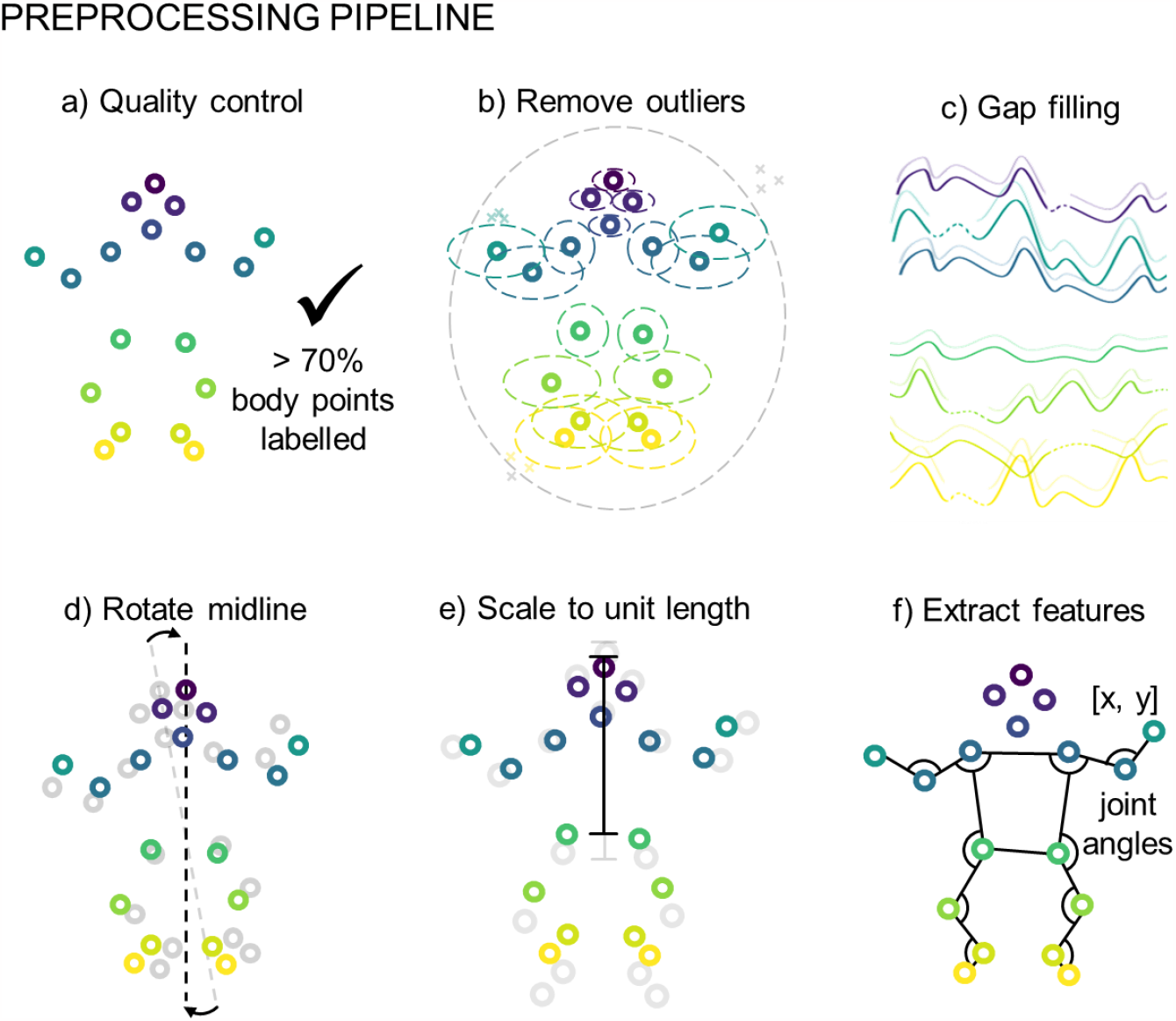
Preprocessing piepline. **a**. Quality control, videos with less than 70% of body points labelled on average were excluded from further analysis. **b**. Outlier removal, outlier body points were removed (denoted by x) when outside of ellipical envelope for the whole body or each body point indiviudally. **c**. Gap filling using linear interpolation for gaps less than 5 frames, or a multivariate imputer for gaps larger than 5 frames. **d**. Body points were rotated on a frame by frame basis ensuring midline of body is aligned to the vertical. **e**. Body point position was scaled to unit length based on infant size, unit length distance crown to mid hip. **f**. For each frame x- and y-coordiates and additional features consisting of joint angles from the left and right shoulder, elbow, hip, knee and ankle were extracted.

#### Quality control

To ensure high-quality movement data from each video was available for further analysis, we established a quality control measure based on body point labelling. Only videos in which more than 70% of body points were labelled on average across all frames were used. This resulted in the exclusion of 21 videos from further analysis, resulting in a final dataset of 484 videos from 327 infants.

#### Outlier removal

Body point outliers were removed in a two-step process. First, outlying labels were removed using an ellipse envelope centred in the centre of the torso (mid-point of hips and shoulders). The ellipse was scaled relative to infant size, with unit length set to distance between the infant’s crown and hip midpoint. The ellipse was scaled to 3 times unit length in the proximal to distal direction and two times unit length in the medial to lateral direction. Body point labels lying outside of the ellipse were removed. Following this, a similar process was applied to each body point using an ellipse envelope centred at the body point’s framewise median position. Each ellipse was again scaled by unit length with the proximal-distal and medial-lateral scaling set based on observed body point variance from the complete dataset. Body point labels lying outside of their respective ellipses were removed.

#### Gap filling

Where gaps in body point data existed due to missing, removed, or occluded body point labels, linear interpolation was used for gaps of five frames or less. For gaps greater than five frames we used an iterative multivariate imputation^38^, implemented in scikit-learn^39^ (v1.3.0).

#### Adjusting for camera movement

As videos were recorded on hand-held devices, camera movement relative to the infant was apparent during the three-minute video. To account for angular rotations, all points were rotated on a frame-by-frame basis so the mid-line of the body (mid-shoulder to mid-hip), was aligned to the vertical in each frame. In addition, body point position in each frame was normalised to infant unit length, measured as distance from crown to mid-hip.

#### Framerate normalisation

All pre-processed movements data were normalised to the same length. Due to variation in video frame rate, the number of frames in each 3-minute video varied. To account for this, all videos were interpolated to 4500 timepoints in length or a framerate of 25 frames per second using cubic 1D interpolation as needed.

#### Feature extraction

For each frame, we extracted each body point’s *x, y* position in addition to 10 joint angles (left and right shoulders, elbows, hips, knees and ankles; in radians, resulting in *p =* 46 features per frame (*keypoints* × {*x, y*} + *joint angles*).

### Prediction of GMA from movement data

As abnormal general movements can occur at any point during each video, may last for different lengths of time, and occur with different frequencies, we aimed to identify short periods of time where abnormal movements were present in each video and use a sliding window approach to generate subject-level predictions. During model training, each subject’s pre-processed timeseries data was split into short clips of *t =* 128 frames in length (approximately 5 sec.) with *stride =* 8. In each training epoch, we randomly sampled *s =* 1 clip per video, selecting more than one clip per video per epoch did not offer an improvement in model performance and cost more memory and computation (**Supplemental Figure S5**).

#### Model architecture

The model architecture is shown in **Figure 2c**. Each subject’s data is represented as a tensor *S* ∈ ℝ^*s* × *t* × *p*^ where *s* is the number of sampled clips per video, *t* is the number of frames per clip and *p* is the number of features per frame. We employ three 1D convolutions applied along the temporal dimension (*filters =* 64; *kernel size =* 3) with causal padding and *ReLU* activations. After each convolutional layer, we applied batch normalisation (**Figure 2c**). Each convolution was followed by max pooling along the temporal dimension with *window size =* 4 and *stride =* 4. After the final convolution, features of each clip were concatenated across the remaining timesteps to form feature matrix *M* ∈ ℝ^*s* × 128^ (**Figure 2c**). Clip features are then passed through a single fully-connected layer (*units =* 64, *ReLU*). We applied dropout with a rate of 0.5 before and after the connected layer.

To identify features that discriminate subjects with or without abnormal movements, we passed each clip through a sigmoid attention module^40^ (**Figure 2c**). In this context, clips with feature vectors that discriminate between classes are given a larger weight. A clip level context vector, *u*, is assigned to measure the importance of each clip to the final model output. First, each clip, *m*_*c*_ ∈ ℝ^1 × 64^, is passed though a single fully connected layer with weights and bias, *W* and *b*, and a *tanh* activation to create clip level representation, *u*_*c*_:

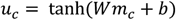

The similarity between each clip’s representation and that of a context vector, *u* is calculated and scaled:

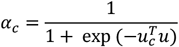

Where *α*_*c*_ ∈ [0,1] and represents the importance of each clip to the final model output. A final representation is calculated though a weighted average of clip features:

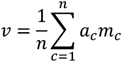

Where *v* is a feature vector representing the sampled clips from each video. The context vector, *u*, and the layer weights and biases are randomly initialised and jointly learned with other model parameters during training. The context vector, *u*, can be considered a ‘signature’ that identifies a discriminative movement within a clip. The resulting weighted outputs form a final feature vector, *v*. We apply a final dropout (0.5) to this vector and pass to a fully connected layer with one unit and *sigmoid* activation to predict the class label of each subject.

We used binary cross entropy (BCE) as the loss function with Stochastic Gradient Descent as the optimiser (Nesterov momentum = 0.9)^41^. As not all randomly sampled clips may contain abnormal movement patterns during each training epoch, we employed label smoothing of 0.1 to account for uncertainty in the assigned sample labels of each batch^42^. The learning rate was set to 0.005, batch size was set to 8 and we added *l*_2_-regularisation of 0.005 to all weight kernels. We trained for a maximum of 10000 epochs, evaluating loss in the validation set and stopping training once validation loss had stopped improving for 100 epochs, retaining the model with minimum loss for testing.

#### Data augmentation

Data augmentation is a common processing step in various image recognition and classification tasks and provides additional protection against overfitting in small sample settings^43,44^. We employed data augmentation methods for timeseries data including random magnitude scaling and time warping^43^ (**Figure 2b**). We used cubic splines to generate a series of random, smooth sinusoidal curves (knots = 3 – 15; mean value = 1.0; sigma = 1.0). During training: i) the timeseries in each clip were multiplied with a randomly generated curve to smoothly scale magnitude across the clip’s length and ii) time warping was applied by smoothly distorting the time interval between points based on another randomly generated curve, shifting the temporal position of adjacent points closer or further apart^43^ (**Figure 2b**).

#### Model calibration and class imbalance

To account for the difference in class frequencies between normal and abnormal GMA (normal = 408 videos; abnormal/absent = 76 videos), we oversampled the minority class by a factor of 5 during training. For each video in the training sample with an abnormal GMA rating we sampled 5 sets of clips during each training epoch, resulting in approximately equal number of training samples from each group.

While resampling methods can improve model performance in imbalanced datasets, they can result in miscalibrated models due to the difference in class frequencies between the original sample population and the oversampled training set^45,46^. We employed Platt scaling^47^ as a post-training method to calibrate model predictions. Model calibration was performed by fitting a logistic regression over model predictions in the validation dataset, the parameters of which are used to transform model outputs to calibrated probabilities at inference.

#### Metadata

Age at video acquisition and birth cohort (extremely preterm or term-born) are both potential confounders that can affect GMA ratings^13^. To incorporate metadata into the model, we applied an additional 1D convolution (*filters =* 4, *kernel size =* 1) to a feature vector of age at video acquisition and categorical group membership (preterm or term-born). The outputs were concatenated with the video features prior to the final layer for classification (**Figure 2c**).

#### Model evaluation

At inference, each test subject’s timeseries data were split into 547 overlapping clips (*t =* 128, *stride =* 8) which were passed with associated metadata through the trained and calibrated model to generate the final model output from the attention-weighted sum of all clips.

To evaluate model performance, we performed cross-validation by splitting the data into three subsets: train (70%), validate (15%) and test (15%), ensuring that the proportion of infants with abnormal movements were similar across subsets and, for infants with more than one video, that both videos were included in the same subset. Model performance was evaluated in the test set using the area under the receiver operating curve (AUC), balanced accuracy (BA), specificity, sensitivity and positive and negative predictive values (PPV; NPV). Cross-validation was repeated 25 times, each with random splits of the dataset. Performance metrics in the test set are reported as average values across the 25 cross-validation repeats. We explore the impact of different parameter choices on model performance in the Supplemental Material (**Figure S5**; **Figure S6**). To examine important model features, we calculated model saliency for each test output using vanilla gradient maps^41^.

#### Baseline model

We compared model performance to alternative models based on logistic regression. For each video, we extracted a set of dynamical features previously shown to perform well in timeseries classification tasks^32^, resulting in *p =* 24 features per timeseries. We concatenated timeseries features for each body point coordinate and joint angle along with associated meta data (age and birth cohort) into a single feature vector of length = 1014. Using this data, we trained an *l*_2_-regularised logistic regression model to predict GMA. As with the convolutional model, we performed 25 cross-validation repeats, splitting the data into 85% training and 15% testing sets. Regularisation strength was set using a grid search (10^−3^ – 10^3^) in a nested 5-fold cross-validation of the training data. To enable additional flexibility in the model, we also implemented a nonlinear kernelised logistic regression using Nystroem kernel approximation^48^. The baseline models were implemented in *scikit-learn*^39^ (1.0.2), timeseries features were extracted using *pycatch22*^32^ (0.4.2).

#### GMA prediction and development at 2 years

Participants were followed up at 2-years’ corrected age and their development assessed using the Bayley Scales of Infant and Toddler Development-3^rd^ edition (Bayley-III) for motor, cognitive and language domains. Bayley-III scores were available for 292/327 infants (441 infant videos) for motor and cognitive domains and 262/327 infants (400 infant videos) for the language domain^13^. Each video was assigned a single GMA prediction label based on the GMA prediction label most frequently assigned during the 25-fold cross validation. This was done for each variant of model metadata inputs: movement data only (none), birth cohort, age at acquisition and combined birth and age (both). For each model variant we compared 2-year outcomes between GMA-prediction groups using an independent two sampled t-test (two-sided). To determine the association of birth cohort and GMA prediction with 2-year outcomes we performed a two-way ANOVA (factors: birth cohort and GMA prediction group, interaction birth cohort*GMA prediction group). As 2-year outcomes are likely confounded by birth cohort, we stratified by birth cohort and performed independent two-sample T-tests (two-sided) between GMA prediction groups.

## Data Availability

Data: The data that supports the findings of this study are available from the corresponding author upon reasonable request. The data is not publicly available due to privacy and ethical restrictions.
Code: We used DeepLabCut to track body points in videos. Code for pre-processing body point data after DeepLabCut, training machine-learning models, analysis of the results are available in our GitHub repository https://github.com/epassmore/infant-movements.

## Data availability

The data that supports the findings of this study are available from the corresponding author upon reasonable request. The data is not publicly available due to privacy and ethical restrictions.

## Code availability

We used DeepLabCut to track body points in videos. Code for pre-processing body point data after DeepLabCut, training machine-learning models, analysis of the results are available in our GitHub repository https://github.com/epassmore/infant-movements.

## Acknowledgements

We would like to acknowledge the parents/families of infants who participated in the study. We would also like to acknowledge the extended Victorian Infant Collaborative Study team for their contribution in collecting infant and 2-year follow up data. We would like to acknowledge funding from the Rebecca L Cooper Medical Research Foundation (PG2019421 to G.B.), National Health and Medical Research Council Investigator Grant (1194497 to G.B.), NVIDIA Corporation Hardware Grant program, The Royal Children’s Hospital Foundation, Melbourne and the Murdoch Children’s Research Institute Clinician Scientist Fellowship.

## Author contributions

Conceptualisation: E.P, G.B and A.S. Data collection: A.K, J.O, A.E. and general movements assessment. E.P and S.G annotated infant videos. Data curation: E.P. A.K and S.G. Data analysis: E.P, S.G and G.B. E.P and G.B wrote the code. J.C and A.S oversaw research study to collect infant videos. G.B oversaw the current study. E.P and G.B wrote the manuscript with input from all authors.

